# Innovating Pathology Learning via Kahoot! Game-based Tool: A Quantitative Study of Students’ Perceptions and Academic Performance

**DOI:** 10.1101/2021.03.06.21253040

**Authors:** Fatma Alzahraa Abdelsalam Elkhamisy, Rita Maher Wassef

## Abstract

**Introduction:** Pathology teaching for medical undergraduate students linking basic and clinical sciences together is a challenging task. Kahoot! is a game-based online digital formative assessment tool that can engage students in its learning. This study analyzed the effect of Kahoot! use on studentś learning of Pathology.

**Methods:** The study was carried out on the first-year Pathology students at Helwan University, Faculty of Medicine, after ending a basic Pathology course. The study is a retrospective quasi-experimental quantitative study. Academic performance of students in Pathology was compared between Kahoot! and non-Kahoot! users (55 students each). In addition, an online survey was introduced to the 55 Kahoot! user students to investigate their perceptions on Kahoot!. Survey and test score data were analyzed by appropriate tests using IBM-SPSS (Statistical Package for Social Sciences) Version 21.0. The level of significance was *P* <0.05.

**Results:** Kahoot! enhanced Pathology understanding (83.6%), retaining knowledge (87.3%), made learning fun and motivating (89.1%). Other mentioned advantages of Kahoot! were practicing for the exam (40%), simple and easy to use (36.4%), competitive (18.2%), self-confidence booster (10.9%), forming a comprehensive image of the lecture (9%), quick (9%), and imagining skills booster (5.5%). Mentioned disadvantages included no explanation for the answers to the questions (20%). A quarter of the students stated that the time limit for the questions was short (27.3%). Kahoot! use was significantly associated with better Pathology academic performance (P=0.001), and it was not related to the general academic performance of the students (P=0.06). The majority of users (85.4%) recommended its continuous future use.

**Conclusions:** The study offers an endorsement to the use of Kahoot! for gamifying formative assessment of Pathology and can provide a basis for the design of an online Kahoot!-based continuous formative assessment plans implemented outside-classroom in the Pathology curricula.

## Introduction

Pathology learning requires continuous training of cognition and skills ^[1]^. Continuous formative assessment helps students in learning as it permits learners to demonstrate their development of skills, and receive support and feedback for more improvement.^[2]^ Finding methods that engage and motivate students to be continuously evaluated,^[3]^ and seeking improvement is needed.

Kahoot! is a formative assessment freely available online tool that is based on the concept of game-based learning. Kahoot! creates a competitive environment through providing time limits and scoring, and is supported by mobile devices. It was launched in 2013, ^[4]^ by Mobitroll, which is a collaboration between the Norwegian University of Science and Technology with the British company We Are Human.^[5,6]^ Kahoot! quizzes can be accessed on any appliance that has an internet signal with no location limits. It can be utilized inside or outside the classroom for individual studentś asynchronous practice or synchronous studentś challenges.^[4,7-9]^

Quizzes can be created freely on the platform by create quiz, then selecting the type of the question (MCQ, or true/false), writing it, followed by choosing the assigned time in seconds for each question. The platform asks you to mark the correct answer to be presented to the students when they answer each question as a feedback. Quizzes can be assigned either as an individual practicing tool where each student pracrices asynchronously on the quiz any time they want with no time limits, or as live challenges inside the classroom, or as a timed-challenges between students outside the classroom. In the challenges, quizzes are open only for a certain time limit which represents a window for students who want to compete with each others to answer and a winnerś list according to scores & time taken is generated. ^[8,9]^ To share a quiz, Kahoot! tool auto-generates a sharable code or link for the instructors. Students can access the game by using the Kahoot! application or by browsing the website via a laptop or smartphone. They enter the shared code or browse the shared link and sign up/in and the Kahoot! game starts. The students collect points based on their speed in offering correct answers. ^[4,10]^ Game elements present in Kahoot! quizzes include stimulating music, colorful animations, and a countdown for each question which maintain participants engagement and creates a sense of competition. Kahoot! creates a bar graph for each quiz that shows how many participants chose each of the answers provided, which gives feedback for both students and instructors.^[11]^ Literature reported some advantages as well as challenges that faced Kahoot! student-users in varying educational settings. ^[12]^

The pilot study by Neureiter et al. (2020) is the only research done evaluating the use of Kahoot! in Pathology learning.^[13]^ It evaluated the general studentś perception on the use of Kahoot! and revealed good acceptance of it. It also evaluated the academic performance through pre and post kahoot tests. However, no comparative group was included in the study to evaluate if the increased performance is real compared to a non kahoot using group. Also the pre and post tests and the kahoot use were done all in the same teaching session which makes the evaluation is based on the short term memory of students retaining the acquired knowledge in the classroom.

Our research aims to investigate the effect of using Kahoot! outside classroom in Pathology learning (i.e. studentś perception, and academic performance using a comparative group).

## Materials and Methods

### The setting

The study was carried out on first-year Pathology medical school students at Faculty of Medicine, Helwan University. The school adopts the SPICES curriculum,^[14]^ which has two phases – Phase I consists of Year 1 and 2 (pre-clinical basic medical sciences phase) and Phase II consists of Year 3 to 5 (clinical phase).

Pathology is a part of the multidisciplinary integrated curriculum of the 1^st^ year, starting from the first module. The Pathology course consisted of 10 lectures and 4 laboratory sessions. Its learning took place between September 2019 and February 2020. The course covered most of the basic pathological principles of diseases (adaptation, cell injury, inflammation, healing, repair, and circulatory disturbances).

### Study design

This pilot study employed a retrospective quasi-experimental design. The academic performance of students was analyzed in the two student groups of the study (Kahoot! formative assessment tool users versus non-Kahoot! users). A survey was introduced to the Kahoot! tool users to assess their perception on the tool (Figure 1).

**Figure 1:**
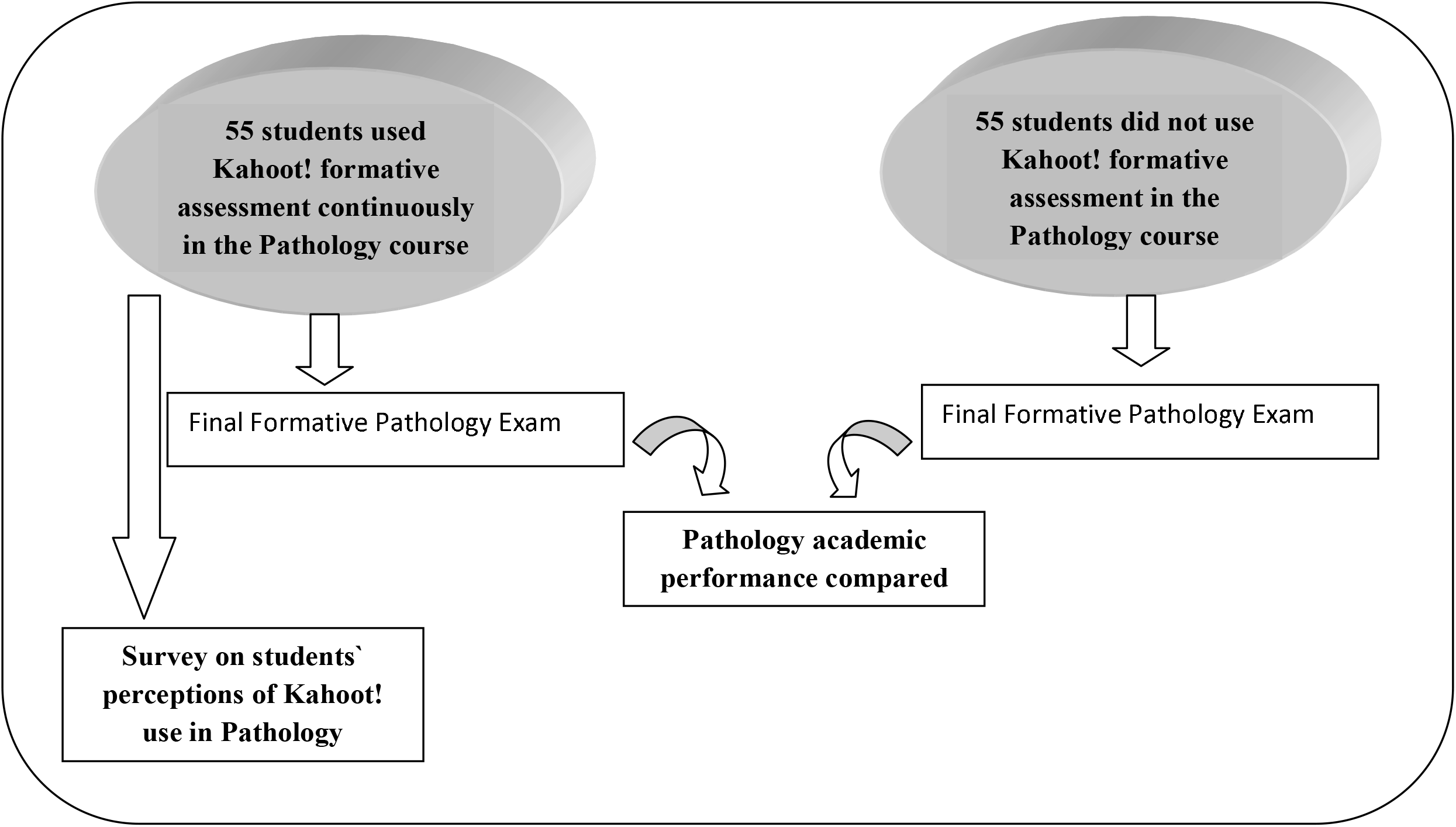
A diagram illustrates the study design.

Before dissemination to students, the survey was sent to two experts in medical education for revision and validation, and slight modifications were done based on their opinions. The final-version survey questions were internally consistent with a calculated Cronbach’s alpha of 0.89.

### Sample

Convenience sample was done; both Kahoot! users and non users from Pathology students were recruited according to their agreement to participate in the survey and in a formative final comprehensive exam. One hundred ten students participated with 55 in each group [users versus non users], no randomization was done for the student groups.

### Implementation

Kahoot! quizzes were used after each lecture over 2 consecutive modules. To be able to participate in the game, invitationś links were shared on the students’ learning management system. There were 2 types of assessments: time-limited individual synchronous challenges, and asynchronous individual practicing quizzes. Kahoot! use as a way of self-monitoring of learning was not compulsory. Each challenge/quiz included 10-25 multiple choice (MCQ) and true/false questions.

The first 3 winners were displayed in descending order by the application at the end of each challenge, and announced on the class’ learning management system (LMS) as Kahoot! challenge champions in Pathology. At the end of the module, the 1^st^ top winner (with total module highest scores) was awarded the Kahoot! champion cup of the module in a celebration attended by all students in the lecture hall.

Google Forms tool was utilized to introduce a survey at the end of the course (2 modules). Students were invited to participate in the online survey shared on their LMS. The survey consisted of a descriptive 21 item questionnaire describing the studentś perceived impact of implementing Kahoot! tool during basic Pathology learning (Supplementary #1). Some questions were adapated from Ismail & Mohammad (2017). ^[2]^ Responses were on a five-point Likert scale with 1 (strongly disagree) to 5 (strongly agree).^[15]^ Adding open-ended written feedback was optional for which breakdown analysis was done to analyze the written data.

### Data Collection

The survey data and studentś test scores were collected on Excel spreadsheets.

### Data analysis

Survey and test score data were analyzed using IBM-SPSS (Statistical Package for Social Sciences) Version 21.0. A Chi-square test was used to compare qualitative variables, and t-tests were used to compare quantitative variables. Z-tests were used to compare proportions. Cronbach alpha test was used to analyze internal consistency of the survey items and a value > is considered internally consistent. The level of significance was *P* <0.05.

### Ethical approval

The study complied with the Helwan University Ethics Committee Guidelines for research with humans and has been approved by the Helwan Medical School Ethics committee (Serial number: 15-2019), organized, and operated according to the declaration of Helsinki. Consent of participation was obtained from all participants prior to their participation in the study as well as for publication of the results.

## Results

This study was conducted on a cohort of 110 first-year preclinical medical students with age range between 17 and 19 years. Participants’ characters are shown in table 1

**Table 1:**
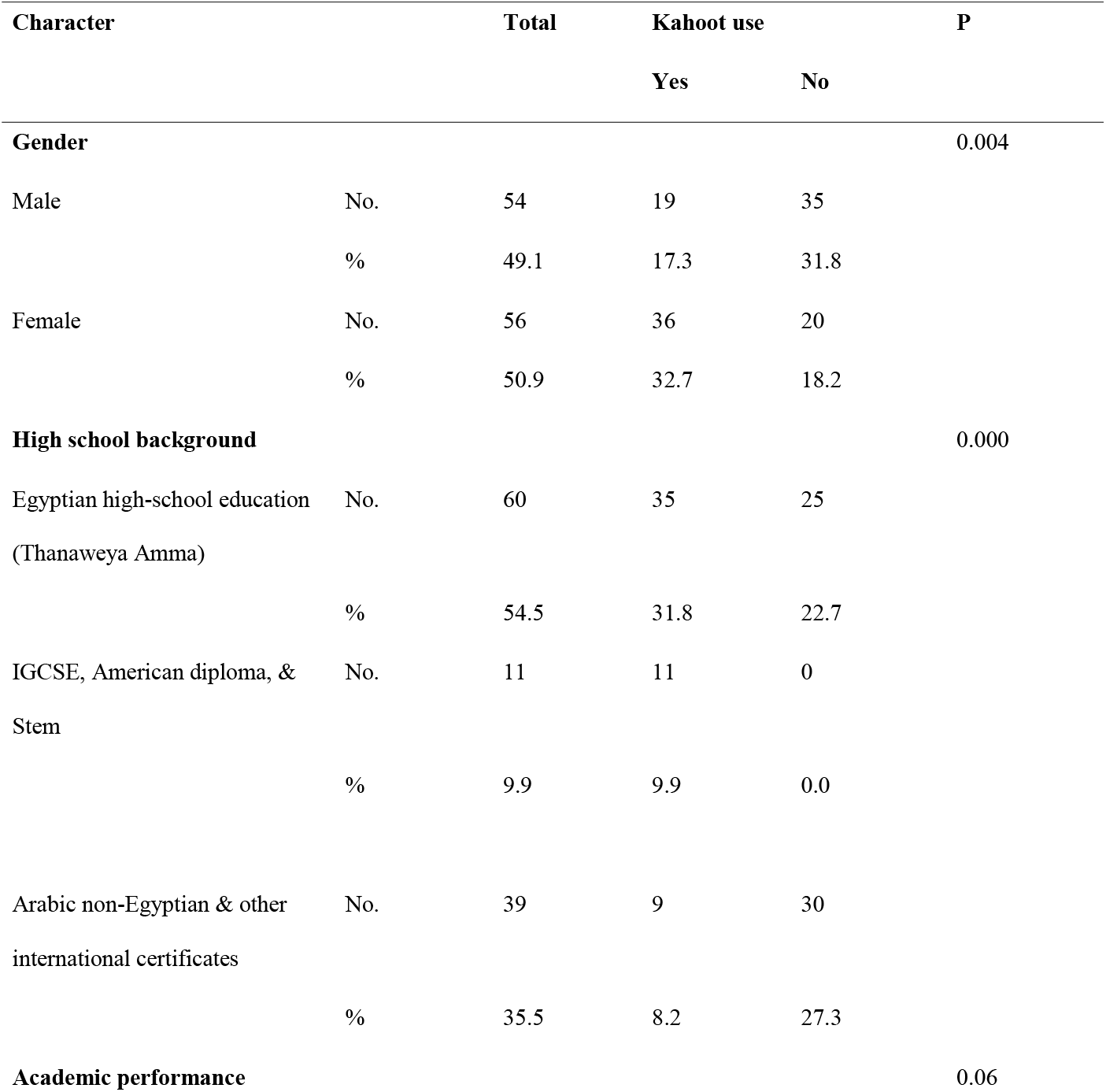
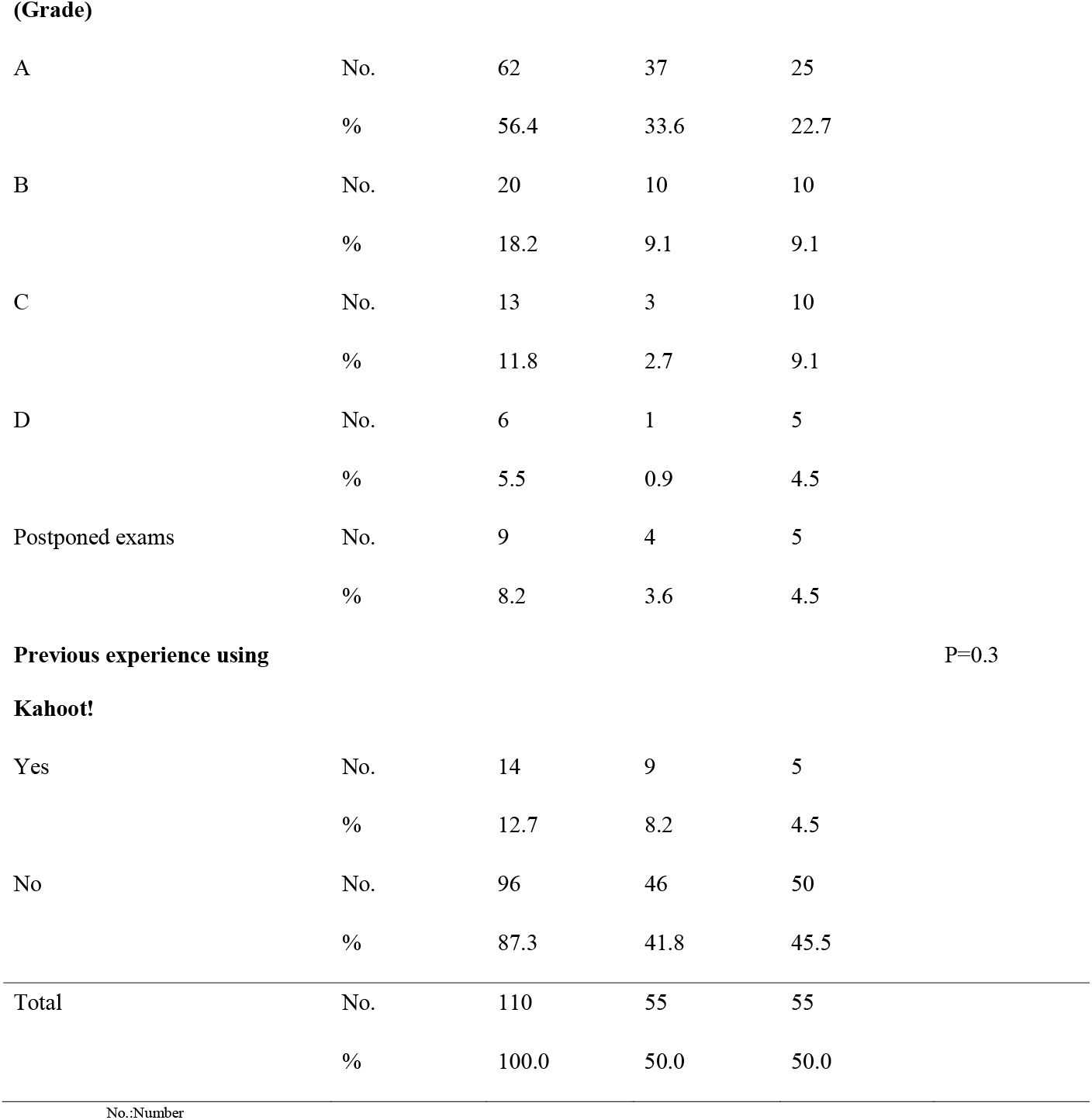
Criteria of Kahoot! versus non Kahoot! users among participants of the study.

### Analysis of Criteria of Kahoot! tool users

Female students who used the Kahoot! tool were significantly higher than male students (*P* 0.004). The type of high school education system also significantly affected it (*P* 0.000). Z tests showed a high difference between students of stem, American diploma, and IGCSE systems who all used Kahoot!, compared to students of Arabic and other international high school systems who used it less. Only 8.2% of students used Kahoot! and had a previous experience on using it (n=9). Previous Kahoot! use did not significantly affect its re-use in Pathology (*P* 0.3). (Table 1)

### Analysis of the academic performance of students using Kahoot!

Regarding the Pathology academic performance, scores of students who used Kahoot! tool were significantly higher (*P* 0.001) in the comprehensive formative Pathology exam held at the end of the module (mean 15.04, SD ±6.15) compared to those who did not (mean=11.00, SD ±5.82).

Regarding the general academic performance of the students, it was described based on participantś grades in the end module summative integrated exam held during the research duration. Most Kahoot! users in the study had grade A, followed by grade B. It was not significantly related to Kahoot! tool use (*P* 0.06). (Table 1)

### Analysis of studentś preferences and perceptions of using Kahoot! as a formative assessment tool

Of the 55 students who used Kahoot! tool; 72.7% (n=40) preferred the individual practicing asynchronous quizzes over the synchronous studentś challenges (n=15, 27.3%). Overall, the majority of students perceived Kahoot! as a reason for increasing their interest in Pathology studying (n=51, 92.7%), beneficial for its learning (n=48, 77.3%), and enhancer for its understanding (n=44, 83.6%). The majority (85.4%) recommended its continuous future use in learning Pathology (n=47). This is explained by the following positive extracted results:

Over 90% of students (n=50, 90.9%) considered it helpful to focus on the subject details, retain knowledge (n=48, 87.3%), and correct misconceptions on the subject (n=45, 81.8%), as well as being an effective method for reflective learning (n=46, 83.7%). The majority (89.1%) of users perceived Kahoot! as an effective method to provide feedback (n=49). It motivated users for learning (n=45, 81.9%), and made their learning experience fun (n=49, 89.1%). (Table 2, Figure 2)

**Table 2:**
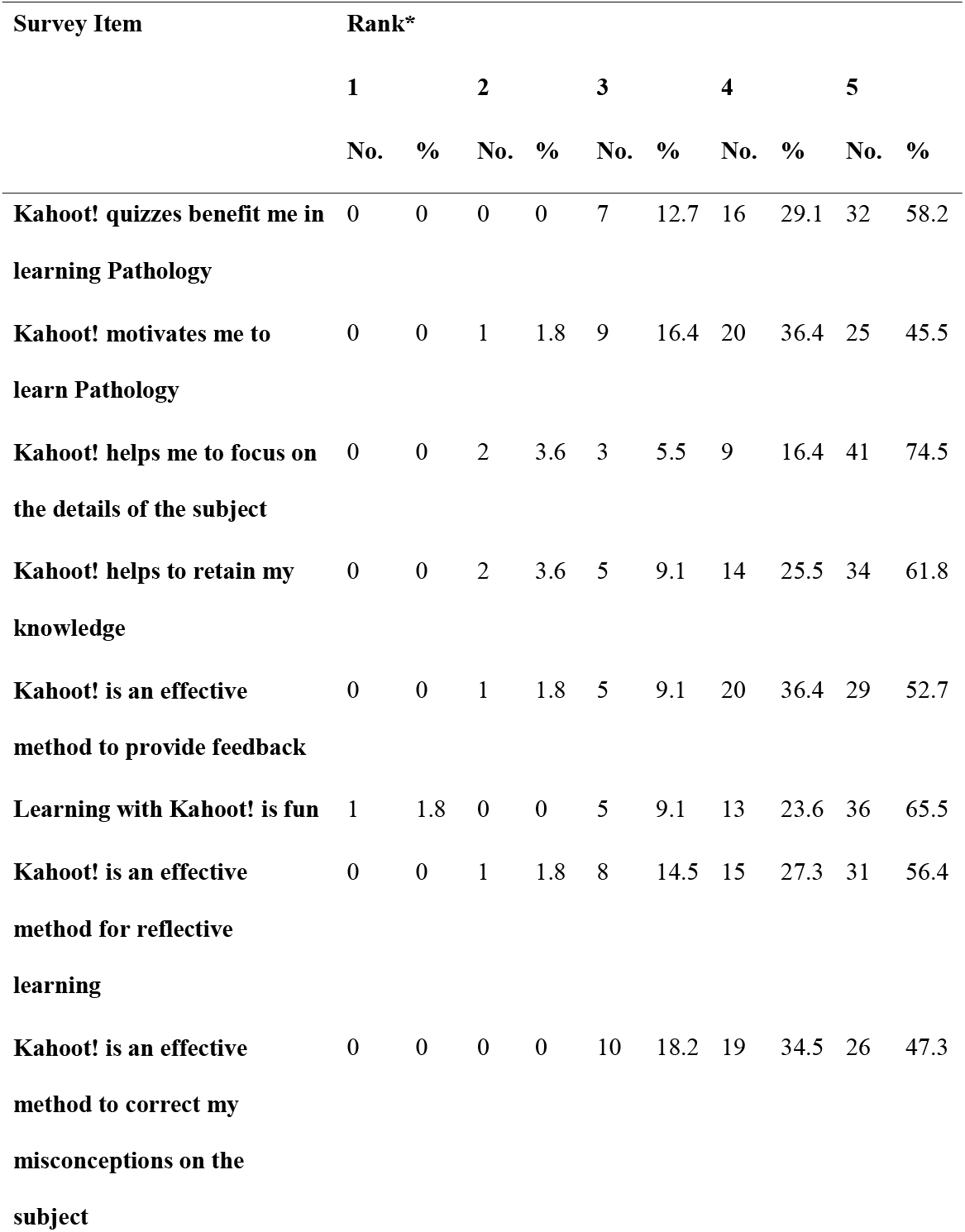
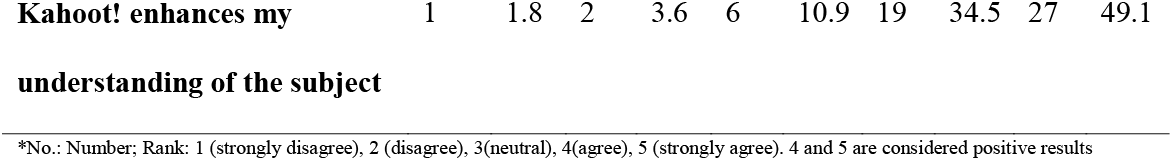
Studentś perceptions of Kahoot! Use.

**Figure 2:**
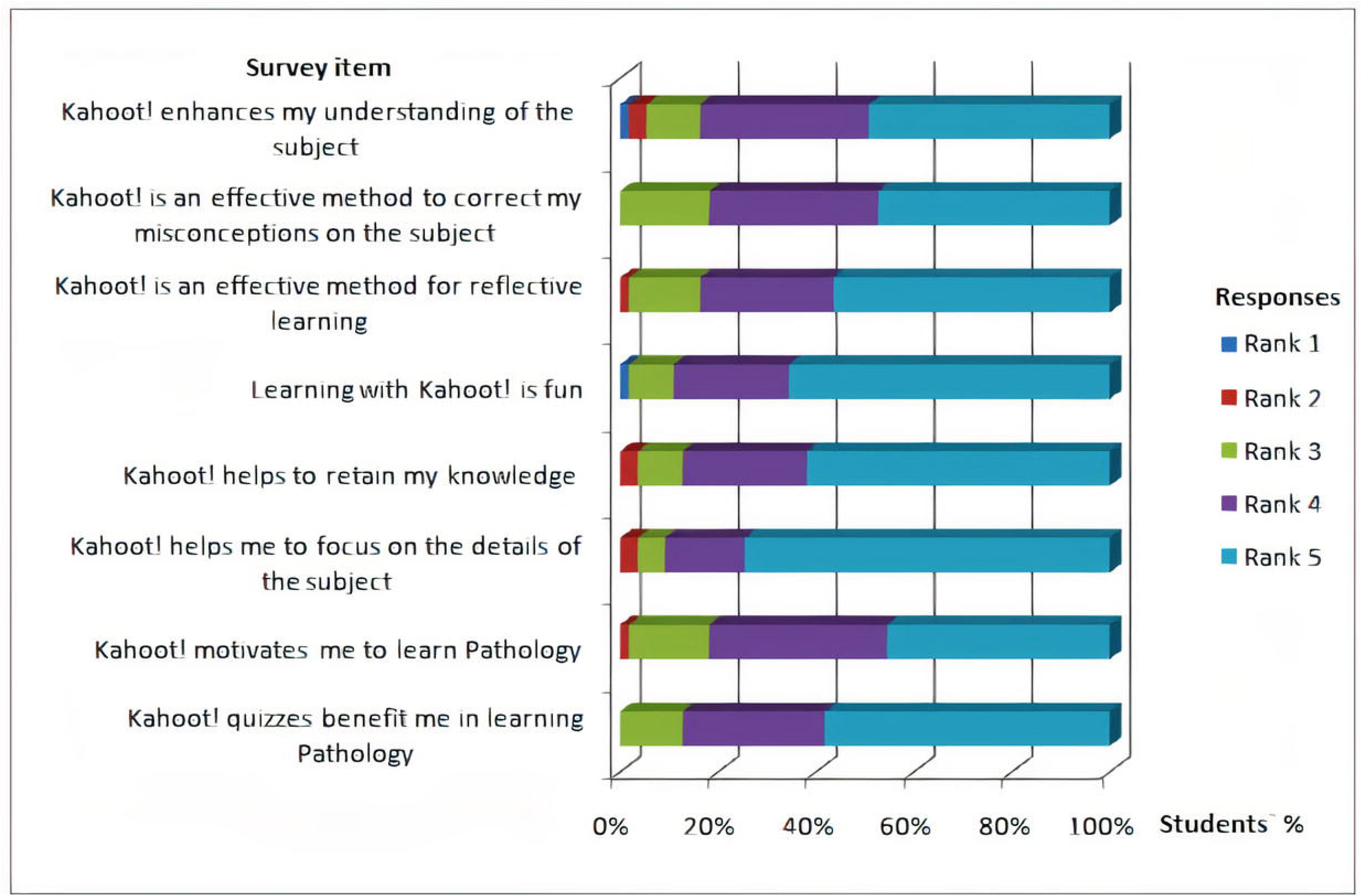
A chart illustr ates studentś perceptions of Kahoot! use.

Kahoot! use is considered a useful tool for assessment of higher cognitive skills by majority of users; analysis (problem solving skills) (n=38, 69.1%), evaluation skills (n=34, 61.8%), application on new situations (n=51, 92.7%), and understanding (interpretation skills) (n=38, 69.1%). (Table 3, Figure 3)

**Table 3:**
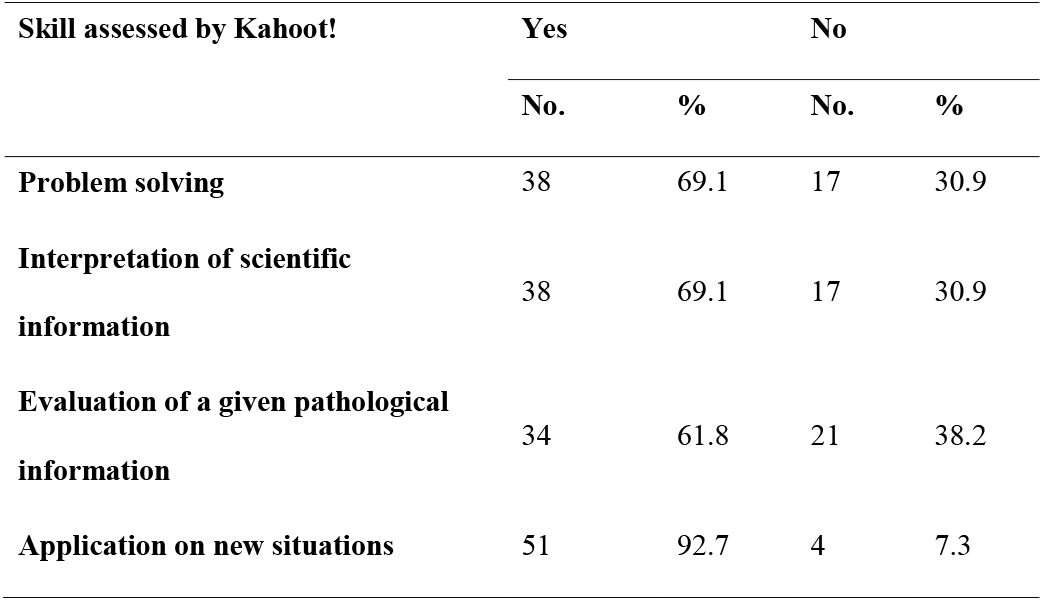
Studentś perception of the level of cognition assessed by Kahoot! use.

| **Skill assessed by Kahoot!** | **Yes** |  | **No** |  |
| --- | --- | --- | --- | --- |
|  | **No.** | **%** | **No.** | **%** |
| **Problem solving** | 38 | 69.1 | 17 | 30.9 |
| **Interpretation of scientific information** | 38 | 69.1 | 17 | 30.9 |
| **Evaluation of a given pathological information** | 34 | 61.8 | 21 | 38.2 |
| **Application on new situations** | 51 | 92.7 | 4 | 7.3 |

**Figure 3:**
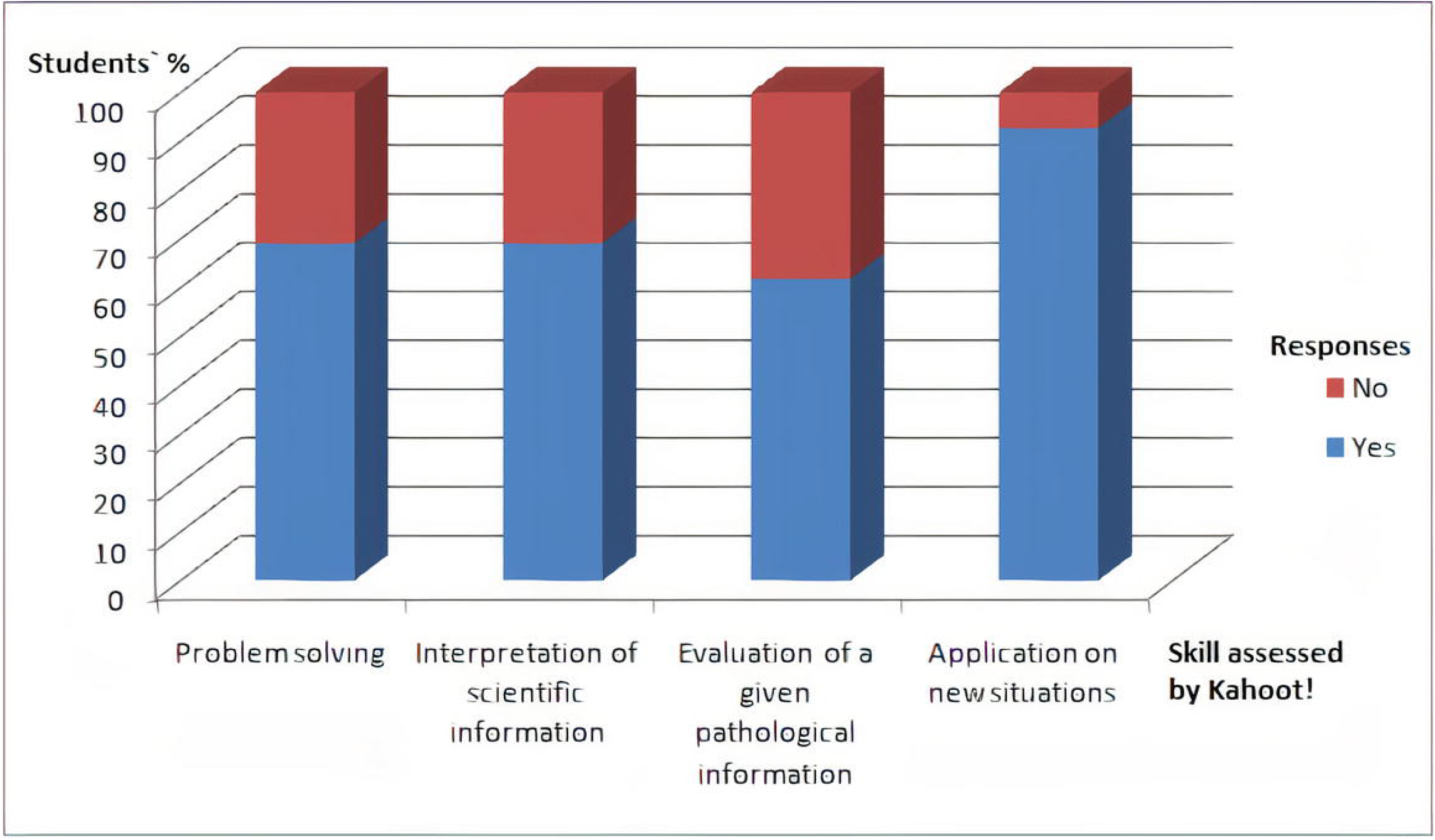
A chart illustrates studentś perception of the level of cognition assessed by Kahoot! Use.

Other mentioned advantages of Kahoot! in a descending order were practicing for exam (n=22, 40%), simple and easy to use (n=20, 36.4%), competitive (n=10, 18.2%), self confidence booster (n=6, 10.9%), forming a comprehensive image of the lecture (n=5, 9%), quick (n=5, 9%), and imagining skills booster (n=3, 5.5%). Mentioned disadvantages included no explanation for the answers of questions (n=11, 20%). A quarter of the students stated that the time limit for the questions was short (n=15, 27.3%).

## Discussion

Our study results showed that using Kahoot! as the game-based tool in learning Pathology is well perceived by students as a learning booster and engager, and increased their academic performance in this medical science.

Medical students in the pre-clinical years may be upset by the amount of knowledge they need to gain, and educators are often faced with the challenging responsibility of teaching a large volume of content in a short time frame.^[16]^ The use of innovative methods in basic science teaching is mandatory nowadays to tackle this problem while keeping up with the new generation of learners. Studies have suggested that today’s students tend to stay more engaged in the educational activities with technology involved.^[17, 18]^

Pathology as a basic medical science linking academic knowledge to clinical cases has always been considered challenging, as students are required to understand, interpret, apply, evaluate, problem solve and memorize a large sum of information about the human body. Innovations in teaching Pathology such as virtual microscopy, public engagement, and podcasts have been tried over the past few years.^[19, 20]^

Games are helpful to obtain high academic performance, motivation, and improved classroom dynamics. Incorporating games in learning started to expand in higher education. Games may be used to overcome some limitations of the traditional face-to-face teaching.^[11]^ Besides, the game-based learning theory is based on that commitment in performing a task while playing stimulates the higher functions of the brain for active learning; this increases knowledge retention by helping the brain transform information from short to long-term memory.^[21]^

Integrating a game-based tool, like Kahoot!, in learning can allow students to revise and apply what they have learned in an interactive, fun, and competitive way.^[7]^ Enhancing game-based learning can enhance medical studentś engagement with Pathology and the findings of our study are consistent with the literature on digital games.^[22]^ Learning methods and platforms that incorporate gamification as Web-based programs, mobile applications, and virtual patient simulations have proven successful in engaging students in learning activities.

To elicit students’ interest and develop their skills, continuous formative assessments in the form of online competitions using Kahoot! were carried out in this study. Besides, our first-year preclinical medical students were challenged with one new Kahoot! competition after each lecture over 2 consecutive Basic Sciences modules to boost their learning by permitting the immediate application of their acquired new knowledge.^[23]^

In our study, out of a cohort of 110 students; fifty five with different high school types and educational systems used the Kahoot! game-based tool. There was a significant association between the high school type of students and their use of the Kahoot! tool in the faculty. All students of stem, American diploma, and IGCSE high school systems used it.They were followed by students of other systems; Egyptian high-school education (Thanaweyya Amma), followed by other non-Egyptian Arabic certificates. So, the type of learning received in high school impacts studentś choices of learning methods in higher education. However, participation in interactive learning and providing feedback in higher education institutions is not limited to a certain background.^[24]^ More encouragement to try gamification in learning and attention should be given to international students of other Arabic high school backgrounds being the least using group.

Female users in our study significantly exceeded males. Females were also found to favor flexible and interactive e-learning methods by other researchers.^[25]^

The majority of Kahoot ! user students involved in the study never used this application before. Students are willing to try new educational tools offered to improve their learning experience. Thirty six percent of Kahoot! user students reported it is easy to use in their written feedback which might be helpful for students with no previous experience using it.

Using Kahoot!, most users preferred the individual asynchronous practicing quizzes over the synchronous individual challenges. Students need to set their pace for training.

The majority of Kahoot! users appreciated it as a tool for formative assessment. They recognized it as beneficial, facilitating and enhancing learning, and increasing their interest in Pathology studying. Moreover, they recommended its continuous future use in learning Pathology. They found it helpful to focus on and retain scientific details; with correction of misconception. Similar results were obtained by former studies on other basic medical sciences.^[26, 27]^

Furthermore, using Kahoot! was associated with good academic performance in Pathology that was not related to the general academic performance of the students. Scores obtained in the comprehensive formative Pathology exam showed significantly higher performance of Kahoot! tool users. This goes in agreement with Tóth, *et al*.^[9]^ who found that lack of engagement in activities is associated with significantly lower performance. Felszeghy *et al*.^[4]^ reported higher academic performance in the Kahoot! user students in Histology, however, it was not statistically significant. Our results are encouraging and support the use of this tool in the somewhat difficult-nature subject.

Critical thinking, problem-solving, reflectivity, analysis, and contextual learning are cognitive competencies needed for an efficient graduate medical student, and they can be developed starting from preclinical years.^[28]^ The Kahoot! application allows the creation of various types of questions; students agreed that they could practice questions that stimulated their higher cognitive functions. Therefore, not only adding a new educational tool is enough, but choosing how to use it according to the nature of the subject/discipline and the target outcomes is an important factor.

This study confirms the positive effect on academic performance found by Neureiter et al. (2020) in Pathology learning. ^[13]^ On contrary to using the tool inside the classroom just after the lecture; our students used the tool outside classroom so the tool is beneficial on the intermediate and long term memory levels. Our study also used a comparative group to confirm the positive effect of Kahoot! which was not used by Neureiter et al. (2020).

Students considered Kahoot! more competitive and challenging in the open-ended questions of the survey; described it as fun, non-boring, and play-like with attractive better design. Some reported that it boosted their self confidence related to answering exams. However, nearly quarter had concerns about the time limit for each question in the quizzes. Challenges reported in the literature that faced some Kahoot! include technical problems, stress related to not having enough time to answer, and fear of losing (Wang and Tahir, 2020). ^[12]^ Challenges to incorporate new technological means should be considered before applying any tool, however, this should not restrain its trial. The time limit for each question during the quizz design should be taken into consideration and prolonged a little bit than that considered suitable by the instructor to allow students for thinking and training. Instructors may add a 2^nd^ version for each quizz to the platform with the considered “suitable” time assigned for the questions for a 2^nd^ phase training.

Study limitations include small number of participants, being implemented only in a general Pathology course with no systemic pathology included, and being carried out on one batch and one instituion.

## Conclusion

This study assessed the criteria and effects of using Kahoot! as an online game-based tool for continuous outside-classroom formative assessment, on undergraduate preclinical students’ engagement and satisfaction while learning a medical Pathology course. The results offer an endorsement to the use of Kahoot! gamification in Pathology learning and can provide a basis for the design a Kahoot!-based formative assessment plans in Pathology curricula. The additional competition aspect of this tool added a supplementary educational value. Besides, through its flexible question design, it offered the opportunity to stimulate and train studentś higher cognitive skills needed for future practitionerś competency.

## Data Availability

The datasets generated and analyzed during the current study are available at the corresponding author upon request.

## Acknowledgement

The authors would like to thank Prof. Azza H. Zidan, Pathology Department, Faculty of Medicine, Port Said University for her assistance in article revision and editing.

## References

1. Knollmann-Ritschel BEC, Regula DP, Borowitz MJ, Conran R, Prystowsky MB. Pathology Competencies for Medical Education and Educational Cases. Acad Pathol. 2017; 4. Article No. 2374289517715040. https://doi.org/10.1177/2374289517715040

2. Ismail MA-A, Mohammad JA-M. Kahoot: a promising tool for formative assessment in medical education. Educ Med J. 2017; 9(2):19–26. https://doi.org/10.21315/eimj2017.9.2.2

3. Wassef R, Elkhamisy F. Evaluation of a web-based learning management platform and formative assessment tools for a Medical Parasitology undergraduate course. PUJ. 2020; 13(2). https://doi.org/10.21608/puj.2020.29543.1070

4. Felszeghy S, Pasonen-Seppänen S, Koskela A, Nieminen P, Härkönen K, Paldanius KM et al. Using online game-based platforms to improve student performance and engagement in histology teaching. BMC Med Educ. 2019; 19(273). https://doi.org/10.1186/s12909-019-1701-0

5. Bicen H, Kocakoyun S. Determination of university students’ most preferred mobile application for gamification.. World J Educ Tech. 2017; 9(1): 18–23.

6. Plump CM, LaRosa J. Using Kahoot! in the classroom to create engagement and active learning: a game-based technology solution for elearning novices. Manag Teach Rev. 2017; 2(2):1–8. Available from: https://doi.org//10.1177/2379298116689783

7. Ofori E, Abulaila Y, DAl-Kurdi D, Jacob S, Miserendino M, Faridi H. Application of Kahoot! as a Teaching and Learning Tool in PharmD Curriculum. The FASEB J. 2020; 34 (S1). https://doi.org/10.1096/fasebj.2020.34.s1.03176

8. Ismail M A, Ahmad A, Mohammad J A, Fakri N M R M, Zarawi Mat Nor M, Najib Mat Pa M.s Using Kahoot! as a formative assessment tool in medical education: a phenomenological study. BMC Med Educ. 2019; 19:230. https://doi.org/10.1186/s12909-019-1658-z

9. Tóth Á, Lógó P, Lógó E. The Effect of the Kahoot Quiz on the Student’s Results in the Exam. Polytech Soc Manag Sci. 2019; 27(2):173–9. https://doi.org/10.3311/PPso.12464

10. Basuki Y, Hidayati YN, Trenggalek S, Wiyata Kediri BW, Kediri WH. Kahoot! or Quizizz: the Students’ Perspectives [Internet]. ELLIC. 2019. Available from: https://doi.org/10.4108/eai.27-4-2019.2285331

11. Aktekin N C, Çelebi H, Aktekin M. Let’s Kahoot! Anatomy. Int J Morphol. 2018; 36(2):716–21

12. Wang AI, Tahir R. The effect of using Kahoot! for learning – A literature review. Computers & Education. 2020; 149 (2020): 103818

13. Neureiter, D., Klieser, E., Neumayer, B., Winkelmann, P., Urbas, R., & Kiesslich, T. (2020). Feasibility of Kahoot! as a Real-Time Assessment Tool in (Histo-)pathology Classroom Teaching. Adv Med Educ Pract.,11, 695–705. https://doi.org/10.2147/AMEP.S264821

14. Harden R M, Sowden A, Dunn W R. Educational strategies in curriculum development: the SPICES model. Med Educ. 1984; 18(4):284–97. https://doi.org/10.1111/j.1365-2923.1984.tb01024.x

15. Likert R A. Technique for the measurement of attitudes. Arch Psychol.1932; 22: 5–55.

16. Craig S, Tait N, Boers D, McAndrew D. Review of anatomy education in Australian and New Zealand medical schools. ANZ J Surg. 2010; 80: 212–6. https://doi.org/10.1111/j.1445-2197.2010.05241.x

17. Al-Hariri M T, Al-Hattami A A. Impact of students’ use of technology on their learning achievements in physiology courses at the University of Dammam. J Taibah Univ Med Sci. 2017; 12(1):82–5. https://doi.org/10.1016/j.jtumed.2016.07.004

18. Bergdah N, Nouri J, Fors U. Disengagement, engagement and digital skills in technology-enhanced learning. Educ Inf Technol. 2020; 25: 957–83. https://doi.org/10.1007/s10639-019-09998-w

19. Sagun L, Arias R. Digital Pathology: An Innovative Approach to Medical Education. PJP. 2018; 3: 7–11. https://doi.org/10.21141/PJP.2018.009

20. Mukundu Nagesh N, Chiva Giurca B, Lishman S. Innovating undergraduate pathology education through public engagement. Virchows Arch. 2018; 472: 853–63. https://doi.org/10.1007/s00428-018-2299-z

21. Banikowski A K, Mehring T A. Strategies to enhance memory based on brain research. Focus Except. Child. 1999; 32(2):1–16. https://doi.org/10.17161/fec.v32i2.6772

22. Janssen A, Shaw T, Goodyear P, Kerfoot B P, Bryce D. A little healthy competition: using mixed methods to pilot a team-based digital game for boosting medical student engagement with anatomy and histology content. BMC Med Educ. 2015; 15(173). https://doi.org/10.1186/s12909-015-0455-6

23. Knowles M S. Andragogy in Action: Applying Modern Principles of Adult Education. 1st ed. Jossey Bass. San Francisco CA 1984.

24. Ogg T, Zimdars A, Heath A. Schooling effects on degree performance: A comparison of the predictive validity of aptitude testing and secondary school grades at Oxford University. BERJ. 2013; 35(5):781– 807. https://doi.org/10.1080/01411920903165611

25. Bruestle P, Haubner D, Schinzel B, Holthaus M, Remmele B, Schirmer D. >et al. Doing E-Learning / Doing Gender? Examining the Relationship between Students’ Gender Concepts and E-Learning Technology. 5th European Symposium on Gender & ICT Digital Cultures: Participation - Empowerment – Diversity, March 5 - 7, 2009 - University of Bremen; 2013. http://www.informatik.unibremen.de/soteg/gict2009/proceedings/GICT2009_Adamus.pdf.

26. Kristen Y, Ethan L, Justin F S. Comparison of Traditional and Gamified Student Response Systems in an Undergraduate Human Anatomy Course. HAPS Educ. 2019; 23(1):29–36. https://doi.org/10.21692/haps.2019.001

27. Sumanasekera W, Chase T, Kaven L, Philip H, Travis J, Thimira S. Evaluation of multiple active learning strategies in a pharmacology course. Currents in Pharmacy Teach & Learn. 2020; 12(1):88–94. https://doi.org/10.1016/j.cptl.2019.10.016

28. Schmidt H G, Mamede S. How to improve the teaching of clinical reasoning: a narrative review and a proposal. Med Educ. 2015; 49(10):961– 73. https://doi.org/10.1111/medu.12775

